# Untargeted plasma proteomics and clinical phenotypes in adolescent depression

**DOI:** 10.64898/2026.07.06.26356404

**Authors:** Aino-Kaisa Piironen, Alexey Afonin, Karoliina Kurkinen, Timo A. Lakka, Tommi Tolmunen, Katja M. Kanninen

## Abstract

**Background:** Depressive disorders are among the most common mental disorders, often emerging in adolescence. Despite advances in biological psychiatry, research on early psychopathology remains scarce. Given the heterogeneity and comorbidity of depressive disorders, identifying biologically informed phenotypes could enhance diagnostic accuracy and personalized treatment approaches.

**Methods:** This study utilized baseline and 6-month follow-up plasma samples (n=47) and clinical data (n=103) of adolescent outpatients with depression (DD, aged 14-19) from the Finnish SMART study and healthy control samples (HC, n=53, aged 15-16) from the Finnish PANIC study. Fasting plasma samples were analyzed using untargeted liquid chromatography-tandem mass spectrometry for proteomics. Data analyses included dimensionality reduction, regression models, correlation analysis, functional enrichment, and factor analysis of mixed data with k-means clustering, including 72 symptom-related items, lifestyle, and socioeconomic scales.

**Results:** Among 756 proteins detected in DD and HC, 308 proteins showed notable, significant (adjusted p<0.01 and Log2FC ≥|1|) alterations in depression. These proteins were enriched in stress-response pathways, including complement and coagulation cascades, energy metabolism, the proteasome complex, and growth factor signaling. Additionally, extracellular matrix proteins were altered. Clinical phenotypes were mostly distinguished by symptom severity, bullying victimization and other trauma-related experiences, social relationships, and medication. Improvement in mood over the 6-month follow-up was associated with shifts in proteins involved in extracellular matrix, cytoplasmic vesicles, and complement and coagulation cascades.

**Conclusions:** Together, adolescent depression displays shared plasma proteomic signatures across its clinical phenotypes, and systemic immune dysfunction, oxidative stress, and extracellular matrix are potential targets for biologically informed interventions in depression.

## 1. Introduction

Depressive disorders are among the most common mental disorders globally (GBD 2019 Mental Disorders Collaborators, 2022), often emerging in adolescence and early adulthood (Kessler et al., 2005). The clinical characteristics of adolescent depression differ slightly from those in adults, and include, e.g., persistently depressed mood, sleep and appetite dysregulation, loss of energy, and impaired social functioning (Jaycox et al., 2009; Rice et al., 2019). Adolescent-onset depression is also associated with higher illness severity and lower socioeconomic development in adulthood (Korczak & Goldstein, 2009), thus increasing the socioeconomic burden of the disease.

Prolonged psychosocial stress is a major risk factor for depression (X. Feng et al., 2025). Chronic stress induces systemic changes in immune, neuroendocrine, and metabolic pathways (McEwen, 2017), and molecular and functional effects on neurodevelopment and brain homeostasis (McEwen et al., 2015; Teicher et al., 2016). These include, for instance, brain structural remodeling via glucocorticoids, excitatory neurotransmitters, endocannabinoids, and other mediators, as well as epigenetic modifications. Cellular processes involved in remodeling include cell adhesion and cytoskeletal reformation, which affect synapses and plasticity (McEwen et al., 2015).

In addition to changes in the brain, systemic alterations are consistently recognized in depression, with the strongest evidence for immunometabolic dysregulation and inflammation (Lamers et al., 2016, 2020; Milaneschi et al., 2021) and oxidative stress responses (Palta et al., 2014). Importantly, peripheral changes interact with the brain via organ-brain axes, such as the gut-brain axis, opening the avenue of a complex brain-body framework (X. Feng et al., 2025). Omics, such as proteomics and metabolomics, are state-of-the-art techniques for investigating dynamic physiological states in health and disease. Previous blood omics studies in adolescent depression have identified alterations in platelet activation, calcineurin signaling (Sokolov et al., 2024), inflammatory markers (Yang et al., 2023), energy metabolism, oxidative stress, and polyunsaturated fatty acid metabolism (Chen et al., 2024; Zhou et al., 2019).

Regardless of the advancements in biological psychiatry, research on early psychopathology remains underrepresented. Existing studies are mainly limited to adults, where the chronicity of symptoms, treatment history, and somatic health conditions have a significant confounding effect. Therefore, in this study, we aimed to identify 1) alterations in the plasma proteome of 47 adolescents with depressive disorders (DD) compared to those of 53 healthy controls (HC), 2) biological and phenotypic subtypes of depression, and 3) the link between changes in plasma proteins and clinical symptoms within a 6-month follow-up. Given the high level of heterogeneity, comorbidity, and poor treatment responses in depressive disorders, this biology-informed approach provides potential targets for interventions and future research, showcasing systemic alterations in adolescent depression.

## 2. Methods

### 2.1. Study population

This study utilized plasma samples and data from two Finnish cohort studies: the Systemic Metabolic Alterations Related To different psychiatric disease categories in adolescent outpatients (SMART) (ClinicalTrials.gov, 2024, NCT06262958) and the 8-year follow-up of the Physical Activity and Nutrition in Children (PANIC) study (ClinicalTrials.gov, 2020, NCT01803776) (Table 1, Table A.1).

**Table 1.** Characteristics of individuals.

| *Cohort study (Group)* | **PANIC (HC)** | **SMART baseline (DD0)** | **SMART follow-up (DD6m)** |
| --- | --- | --- | --- |
| Number of individuals with clinical data | 53 | 103 | 75 |
| Age, mean (SD) | 15.8 (0.4) | 16.5 (1.5) | 17.1 (1.6) |
| Girls, n (%) | 25 (47) | 87 (84) | 65 (87) |
| BMI, mean (SD) | 20.6 (2.79) | 23.0 (4.04) | 23.8 (4.30) |
| BDI, mean (SD) | 1.0 (1.47) | 29.5 (8.48) | 23.2 (12.04) |
| Divorced parents, n (%) | 6 (11) | 50 (49) | 30 (40) |
| Sleep >7h/night, n (%) | 52 (98) | 31 (30) | 29 (39) |
| Medication for depression, n (%) | - | 53 (51) | 38 (76) |

HC=healthy controls, DD0=adolescents with depressive disorders at baseline, DD6m=adolescents with depressive disorders at 6-month follow-up, BMI=body mass index, BDI=Beck Depression Inventory.

We analyzed plasma samples from 47 adolescents with DD and 53 HC. In the DD group, baseline plasma samples (DD0) were obtained from 47 adolescents aged 14-19 years, and 6-month follow-up samples (DD6m) were available from 46 adolescents. Additionally, we used clinical data from 103 adolescents at baseline, of whom 75 participated in the 6-month follow-up. At baseline, 51 adolescents were diagnosed with major depressive disorder (MDD, DSM-IV 296.21-296.34), 20 with dysthymic disorder (DSM-IV 300.4), and 32 with both MDD and dysthymia (i.e. double depression), based on the Structured Clinical Interview for DSM-IV (SCID-IV) (First et al., 1997). These diagnostic subgroups and comorbid psychiatric diagnoses based on DSM-IV were included as binary variables in sensitivity analyses. Participants were receiving “treatment-as-usual” during the follow-up.

Altogether, we used cross-sectional plasma samples from 53 HC aged 15-16 years who were selected from the PANIC study based on the following criteria: 1) no psychiatric, neuropsychiatric, or neurological diagnoses, cancer, or other chronic diseases, 2) the total score in the Beck Depression Inventory (BDI) ≤ 12 (no depressive symptoms) (Beck et al., 1988), 3) the total score in Cohen’s stress scale ≤ 26 (no high stress) (Cohen et al., 1983), 4) no regular use of medications, 5) no self-reported frequent pain occurring more than once per week, and 6) below the sex-specific median for self-reported psychological difficulties in the Adolescent Well-being Questionnaire (Ikävalko et al., 2018).

SMART and PANIC studies comply with the Declaration of Helsinki (World Medical Association, 2013). The Research Ethics Committee of the Hospital District of Northern Savo approved the SMART study (238/2017) and the PANIC study before baseline in childhood (69/2006) and before the 8-year follow-up in adolescence (422/2015). All adolescents and caregivers of those younger than 15 years old in the SMART study provided written informed consent. Adolescents in the 8-year follow-up of the PANIC study also provided written informed consent.

### 2.2. Clinical assessment and questionnaires

In the SMART study, a trained research nurse conducted a diagnostic interview (SCID-IV) (First et al., 1997) for all participants to fill in clinical questionnaires, including the 21-item BDI (BDI-1A) (Beck et al., 1988) for depressive symptoms, the Trauma and Distress Scale (TADS) (Patterson et al., 2002) for adverse childhood events, and the Strengths and Difficulties Questionnaire (SDQ) (Goodman, 2001) for emotional, social, and behavioural difficulties in daily life. The complete list of questionnaires and clinical variables used in this study is provided in Table A.2.

Data on medications for the corresponding time were obtained from the patient files. Confirmed data on medication use were available for all in DD0 and for 50 adolescents (67%) in DD6m. We used data on baseline medication for depression (drug-naïve/medicated) in the cross-sectional analysis comparing DD0 and HC, and data on DD-medication over the follow-up (drug-naïve/medicated at any time point) in the longitudinal DD-specific analysis. Body mass index (BMI) was available for 33 adolescents in DD0 (32%) and 15 adolescents in DD6m (20%). It is possible that depressed adolescents refused to be weighed due to heightened sensitivity to body image concerns.

### 2.3. Blood sampling

At the time of research visits, EDTA-plasma samples were collected at Kuopio University Hospital for the SMART study and at the University of Eastern Finland (UEF) for the PANIC study. In both studies, plasma samples were obtained in the morning after overnight fasting, rested for 30 minutes at room temperature, centrifuged at 2500 x g for 10 min, and frozen at -80 °C. Sample collection and storage were conducted by the Eastern Finland Laboratory Centre Joint Authority Enterprise at Kuopio University Hospital in the SMART study and at the Institute of Biomedicine, UEF, in the PANIC study. Sampling protocols are described in detail in Table A.3.

### 2.4. Mass spectrometry

Plasma samples were processed and analyzed for untargeted proteomics at the Turku Proteomics Facility (https://bioscience.fi/) supported by Biocenter Finland. Briefly, samples were depleted from highly abundant plasma proteins using High-Select™ Top14 Abundant Protein Depletion Resin kit (Thermo Scientific, Rockford, USA) according to the manufacturer’s instructions. The proteins of the depleted samples were acetone-precipitated, subjected to in-solution digestion, desalted, and further processed as previously described (Afonin et al., 2024). For data-independent acquisition (DIA), 800 ng of peptides were injected and analyzed in a randomized order. The liquid chromatography-electrospray ionization-tandem mass spectrometry (LC-ESI-MS/MS) analysis was performed on a nanoflow HPLC system (Easy-nLC1200, Thermo Fisher Scientific) coupled to the Orbitrap Exploris 480 mass spectrometer (Thermo Fisher Scientific, Bremen, Germany) equipped with a nano-electrospray ionization source and FAIMS interface. Compensation voltages of -40 V and -60 V were used. The specifics of the instrument and the analysis parameters are described in (Afonin et al., 2024) with the following differences in this study: A 120 min gradient was used to elute peptides (62 min from 5% to 21% solvent B, followed by 48 min from 21% to 36% solvent B and in 5 min from 36% to 100% of solvent B, followed by a 5 min wash stage with solvent B). MS data were acquired automatically by using Thermo Xcalibur 4.6 software (Thermo Fisher Scientific). In the DIA method, a duty cycle contained one full scan (400 -1000 m/z) and 30 DIA MS/MS scans.

### 2.5. Protein identification and quantification

The direct DIA approach was used to identify proteins, and label-free quantification was performed with MaxLFQ. Data were analyzed by Spectronaut software (Biognosys; version 18.7.240325) using protein databases: Homo Sapiens Swiss-Prot reference proteome (UniProtKB/Swiss-Prot release 2024_01) (Bateman et al., 2025), and Universal Protein Contaminant database (Frankenfield et al., 2022), local normalization based on RT-dependent local regression model (Callister et al., 2006), and the other parameters as previously described (Afonin et al., 2024).

### 2.6. Proteomics data pre-processing

Depleted proteins and contaminants were removed from the proteomics data, followed by sample filtering based on the total number of proteins identified per sample (> 950). Signal drift and batch effects were examined according to the pipeline presented by (Čuklina et al., 2021). No drift or batch effects were identified. Missingness was evaluated for each cohort and time point (HC, DD0, DD6m). Only proteins with ≤ 20% missing values per group were included. Additionally, each sample was required to have ≤ 15% missing protein abundances. Next, protein abundances were normalized by sample medians with multiplication of a global median to adjust the scale, and missing values were imputed using the sample minimum method (Liu & Dongre, 2021). Imputed data were log2-transformed, and resulting negative values were replaced by zeros. Data pre-processing was performed in R (R Core Team, 2024) (v.4.4.3).

### 2.7. Statistical and bioinformatic analysis

All statistical analyses and data visualization were performed in R (R Core Team, 2024) (v.4.4.3) and are described in detail below.

#### 2.7.1. Dimensional reduction and clustering

Principal component analysis (PCA) was used to obtain an overview of proteomic profiles. PCA-based feature selection was done using the *irlba* package (Baglama & Reichel, 2005) and protein loadings for the first five principal components (PC1-5). Factor Analysis for Mixed Data (FAMD) was performed using the package FactoMineR (Lê et al., 2008) for clinical data to identify phenotypic patterns among depressive adolescents. Clinical variables with a maximum of 10% of missing values were included, and the remaining missing data were imputed using the medians for continuous and the modes for categorical variables. K-means clustering was performed for phenotypes using individuals’ coordinates in FAMD and for proteomics-based clusters using detected proteins in the DD samples, and the packages *cluster* (Maechler et al., 2025) and *factoextra* (Kassambara & Mundt, 2020). The optimal number of clusters was examined using the total within-cluster sum of squares and gap statistics (Tibshirani et al., 2001).

#### 2.7.2. Regression analyses

Differentially abundant proteins between DD0 and HC and among DD0 clusters were examined using linear regression modelling with the *limma* package (Ritchie et al., 2015) using the functions lmFit, contrasts.fit, and eBayes. The models were adjusted by age, sex, and having medication for depression at baseline (drug-naïve/medicated). Longitudinal protein changes in the DD group were analyzed using a paired-sample design in *limma* and adjusted for age, sex, and DD-medication over the follow-up. Similarly, follow-up data on symptom scores were analyzed using the packages *lme4* (Bates et al., 2015) and the *lmerTest* (Kuznetsova et al., 2017) for linear mixed-effect models (LMM). These included time point, age, sex, and DD-medication use over the follow-up as fixed covariates and individuals as a random effect. Balanced data from 75 adolescents were used in the first LMM, and the second model included 59 adolescents with DD-medication data. Additionally, associations between longitudinal protein changes and the selected symptom scores based on significant findings (Benjamini-Hochberg (BH) adjusted p-value < 0.05) in the LMM models were examined using multiple linear regression. Three models were run and adjusted for baseline protein abundance and a symptom score (Model 1), complemented with age and sex (Model 2), or DD-medication use over the follow-up (Model 3). Proteins that were significant in all three longitudinal models were selected. In all regression models, p-values were corrected for multiple testing using the BH method to control false discovery rate (FDR) (Benjamini & Hochberg, 1995), and adjusted p-values (adj. p) < 0.05 were considered statistically significant.

#### 2.7.3. Functional enrichment and pathway analysis

Highly significantly different proteins (adj. p < 0.01, absolute Log2FC ≥ 1) between DD0 and HC in the baseline linear regression analysis were included in the StringDB (Szklarczyk et al., 2023) functional enrichment and protein-protein interaction analyses. In the follow-up analyses, nominally significant (p < 0.05) proteins were used for enrichment. The complete set of proteins detected in this study (n=3268, excluding depleted proteins and contaminants) was used as a statistical background.

#### 2.7.4. Correlation analysis

PC scores and FAMD coordinates for individuals were correlated using Spearman’s correlation to link proteomics and phenotypic profiles. The first five dimensions in PCA and FAMD were used, and two-sided Spearman’s rank correlation coefficients and BH-adjusted p-values were calculated for each pair.

#### 2.7.5. Sensitivity analysis

Linear regression models for proteomics data were repeated using a group*sex interaction term and contrasts in *limma* to calculate statistics sex-specifically without reducing the total number of samples. Additional regression analyses were performed to consider the possible confounding effects of smoking, alcohol use, and BMI, along with age, sex, and medication use. Data on smoking and alcohol use were available for all adolescents. BMI was included in a separate model using a subset of adolescents (21 DD0 and 49 HC) with reported BMI measures. Furthermore, the PCA plots of DD0 samples were colored by sex, medication status, and the diagnostic subgroups: MDD, dysthymia, double depression, and comorbid psychiatric diagnoses.

## 3. Results

### 3.1. Protein identification

Shotgun proteomics identified 3505 plasma proteins (Table A.4). After removing the depleted proteins and contaminants, 3268 proteins were included in further processing. A higher number of proteins were identified in the DD plasma samples (mean 1555, SD 202) compared to the HC plasma samples (mean 1218, SD 208). Four HC samples were excluded due to a low total number of detected proteins (< 950). After preprocessing, 756 shared proteins across 47 DD0 samples and 49 HC samples were included in regression models. Additionally, 1074 common proteins were included in the analysis of 46 DD0 and 44 DD6m samples that met the preprocessing criteria (see Methods).

### 3.2. Plasma protein alterations in depression

PCA using the baseline samples revealed distinct groups of depressed and healthy adolescents, and the first two PCs explained 48.3% of the variance (Fig. 1A). Linear regression identified 556 significantly (adj. p < 0.05) altered proteins in adolescents with depression (Table A.5a). Among those, 308 most distinct proteins (adj. p < 0.01 and Log2FC ≥ |1|), of which 167 were decreased and 141 increased (Fig. 1B), were included in the enrichment analysis. Most of these 308 proteins remained highly significant (adj. p < 0.01 and Log2FC ≥ |1|) after adjusting for smoking and alcohol use or BMI, resulting in 302 and 281 overlapping proteins, respectively, and 279 common proteins across all three models (Fig. B.1, Table A.5b). Sex-specific analysis revealed more alterations between DD and HC in girls (313 proteins with adj. p < 0.01 and Log2FC ≥ |1|) compared to boys (207 proteins, respectively) (Fig. B.2, Table A.5a).

**Figure 1.**
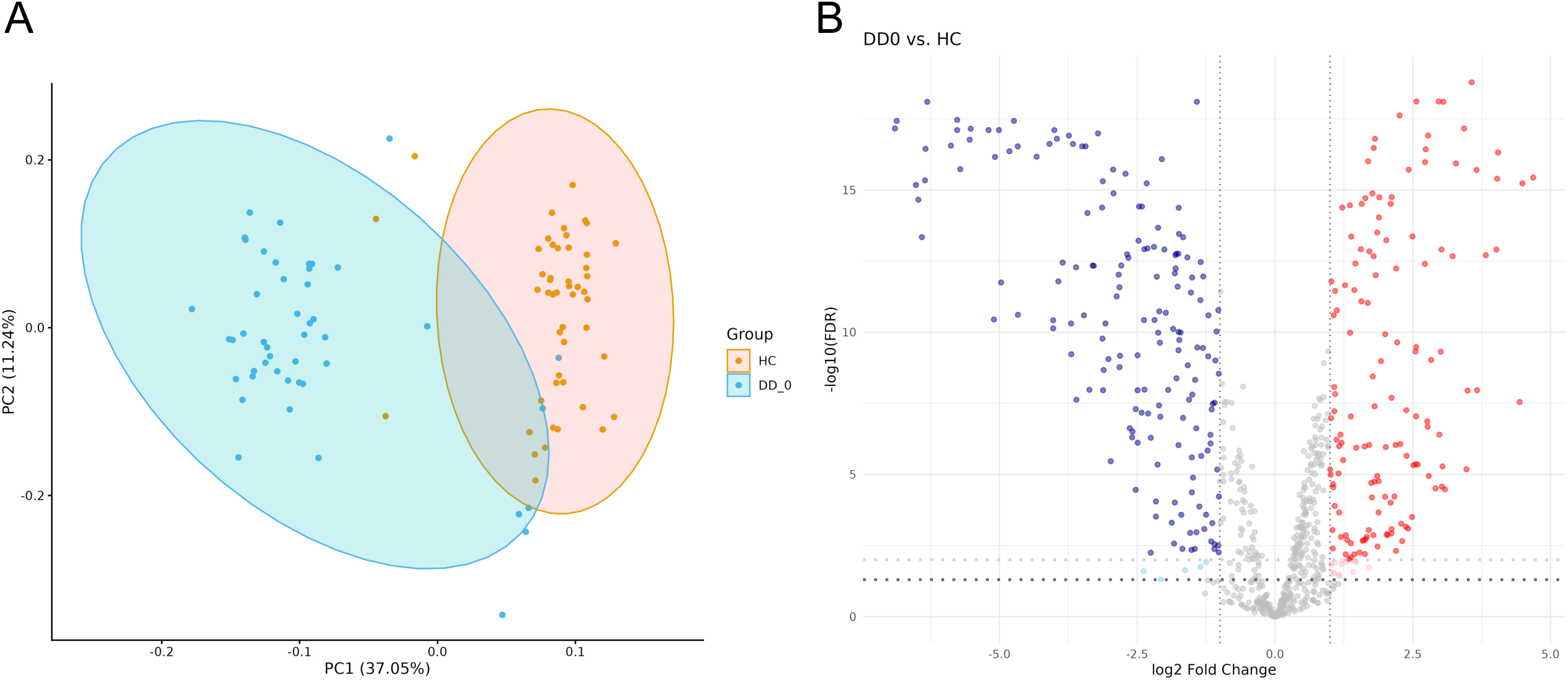
Plasma protein alterations between DD and HC. **A)** PCA for baseline samples. HC: healthy control (n=49), DD_0: adolescents with depression at baseline (n=47), **B)** Volcano plot of 308 significantly altered proteins between DD0 and HC in regression analysis; 167 decreased proteins in blue, and 141 increased proteins in red. Adjusted p-value < 0.01 and the absolute Log2FC ≥ 1, light blue and red indicate additional proteins with adjusted p-value = 0.01-0.05 using the Benjamini-Hochberg method.

Overall, the most distinct proteins between DD and HC were involved in the innate immune system (e.g. complement cascade) and hemostasis, cellular stress responses, and the regulation of insulin-like growth factor (IGF) transport and uptake (Table 2, Table A.6a). Furthermore, mitogen-activated protein kinase (MAPK) family signaling cascades, keratan sulfate biosynthesis, proteasome, KEAP1-NFE2L2 (Kelch-like ECH-associated protein 1-nuclear factor erythroid-derived 2-like 2) pathway, and extracellular matrix (ECM) organization were specifically enriched in girls, whereas boys showed stronger enrichment in glucose metabolism and energy production. Additionally, proteins involved in bacterial respiratory infection and complement protein-related autoimmune disease were enriched in both sexes (Table 2, Table A.6a-c).

**Table 2.** The top enriched pathways between DD and HC.

| Pathway ID | Pathway name | All | Girls | Boys |
| --- | --- | --- | --- | --- |
| HSA-168249 | Innate Immune System | 0.28 ↑↓ | 0.27 ↑↓ | 0.27 ↑↓ |
| HSA-166658 | Complement cascade | 0.64 ↓ | 0.65 ↓ | 0.71 ↓ |
| HSA-109582 | Hemostasis | 0.31 ↑↓ | 0.32 ↑↓ | 0.37 ↑↓ |
| HSA-114608 | Platelet degranulation | 0.52 ↑↓ | 0.54 ↑↓ | 0.62 ↑↓ |
| HSA-381426 | Regulation of IGF transport and uptake by IGFBPs | 0.52 ↓ | 0.51 ↓ | 0.59 ↓ |
| HSA-9711123 | Cellular response to chemical stress | 0.47 ↑ | 0.45 ↑ | 0.46 ↑ |
| hsa05133 | Pertussis (bacterial infection) | 0.64 ↓ | 0.63 ↓ | 0.65 ↓ |
| hsa05322 | Systemic lupus erythematosus | 0.57 ↓ | 0.59 ↓ | 0.62 ↓ |
| HSA-5683057 | MAPK family signaling cascades | 0.31 ↑ | 0.30 ↑ | - |
| HSA-2022854 | Keratan sulfate biosynthesis | - | 0.77 ↓ | - |
| hsa03050 | Proteasome | 0.60 ↑ | 0.59 ↑ | - |
| HSA-9755511 | KEAP1-NFE2L2 pathway | 0.48 ↑ | 0.47 ↑ | - |
| HSA-1474244/<br>CL:19457* | Extracellular matrix organization | -/0.40 ↓ | 0.27/0.44 ↓ | - |
| HSA-70263 | Gluconeogenesis | 0.69 ↑ | - | 0.86 ↑ |
| HSA-70171/<br>CL:8577* | Glycolysis/Glycolysis, and Pentose-phosphate shunt | -/0.64 ↑ | -/0.59 ↑ | 0.65/0.74 ↑ |

Strength and direction of the change (arrows) of the enriched Reactome and KEGG pathways or STRING clusters\*, adj. $p < 0.05$ . Upward arrows indicate that proteins involved in the pathway are predominantly increased in DD0, and downward arrows indicate decreased proteins in DD0, respectively. Double arrows indicate both increased and decreased proteins involved in the pathway in DD0 compared to HC. IGF = Insulin-like Growth Factor, IGFBPs = Insulin-like Growth Factor Binding Proteins, MAPK = mitogen-activated protein kinase, KEAP1-NFE2L2 = Kelch-like ECH-associated protein 1-Nuclear Factor erythroid-derived 2-like 2.

### 3.3. Proteomics clusters in depression

Three proteomics clusters were identified among the DD group at baseline (Fig. 2). The smallest cluster (cluster 3, n=6) consisted of individuals who were mapped closest to HCs in the DD0-HC comparison. Proteomics clusters were not driven by diagnostic subgroups, medication status, or sex (Fig. B.3). Cluster 3, compared to the other clusters, reflected the biological changes identified between DD0 and HC. Additionally, the differentially abundant proteins in cluster 3 were involved in carbon and pyruvate metabolism, amino acid biosynthesis, the citric acid cycle, cell adhesion, and cellular detoxification. However, most changes were significantly enriched only between clusters 2 and 3, whereas those between clusters 1 and 3 were primarily linked to complement and coagulation cascades, hematopoietic cell lineage, and cell adhesion. The most altered proteins between clusters 1 and 2 were involved in EPH-Ephrin signaling, RHO GTPase Effectors, and hemostasis (Table A.7a-c).

**Figure 2.**
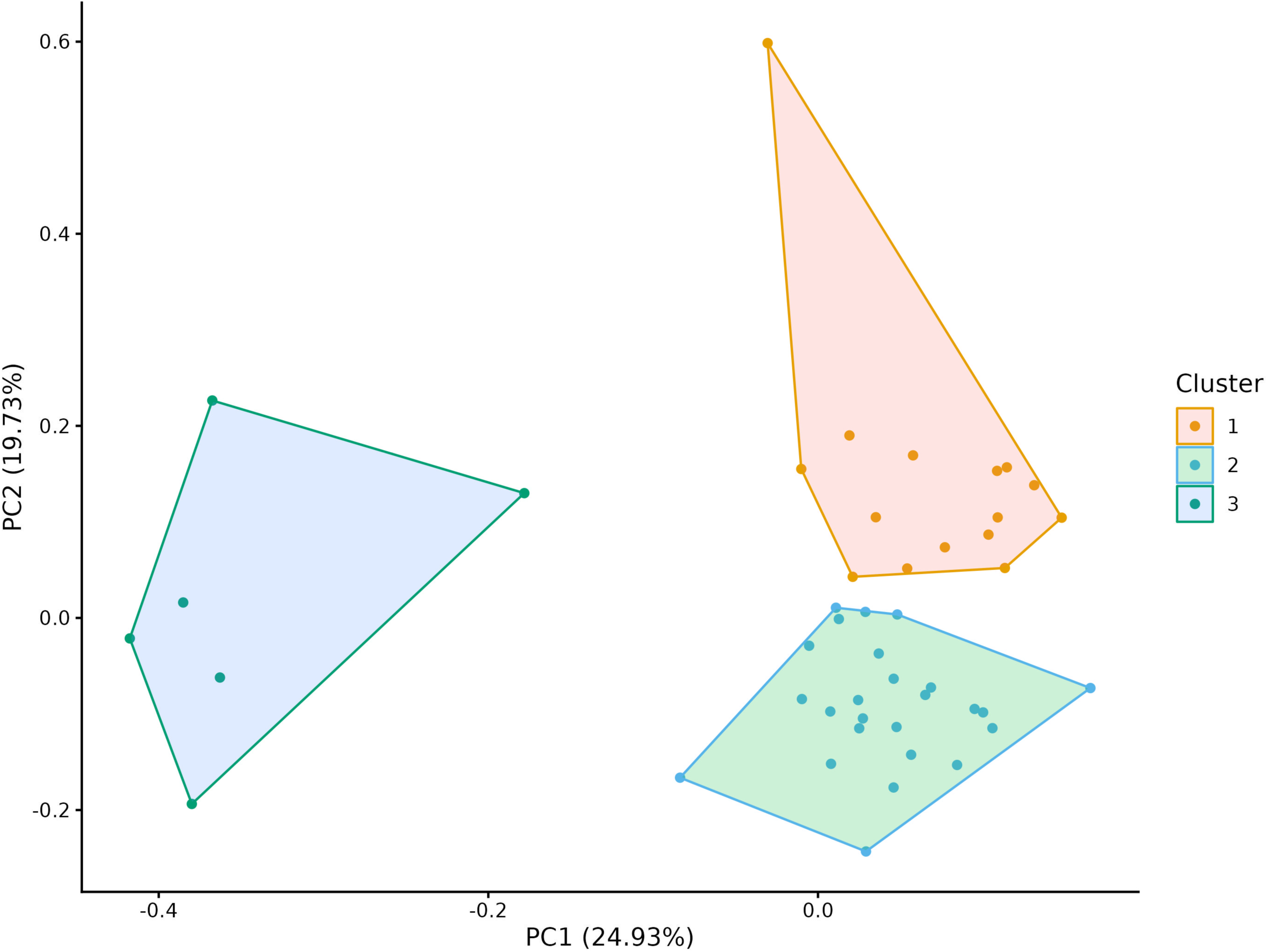
Baseline proteomics clusters of adolescents with depression. Cluster 1 (n=15), cluster 2 (n=25), and cluster 3 (n=6) from k-means clustering. Cluster 3 includes individuals closest to the healthy controls.

### 3.4. Phenotype clusters in depression

Clinical heterogeneity was assessed using Factor Analysis for Mixed Data (FAMD) for 103 adolescents with depression, identifying the baseline symptoms and background variables driving the phenotypic differences. Variables used in the FAMD are listed in Supplementary Table 6. The first two dimensions explained 17.8% of the variance, consisting mostly of internalizing symptoms, bullying victimization and other trauma-related experiences, social relationships, age, medication, sex, and school attendance (Fig. 3A-B).

**Figure 3.**
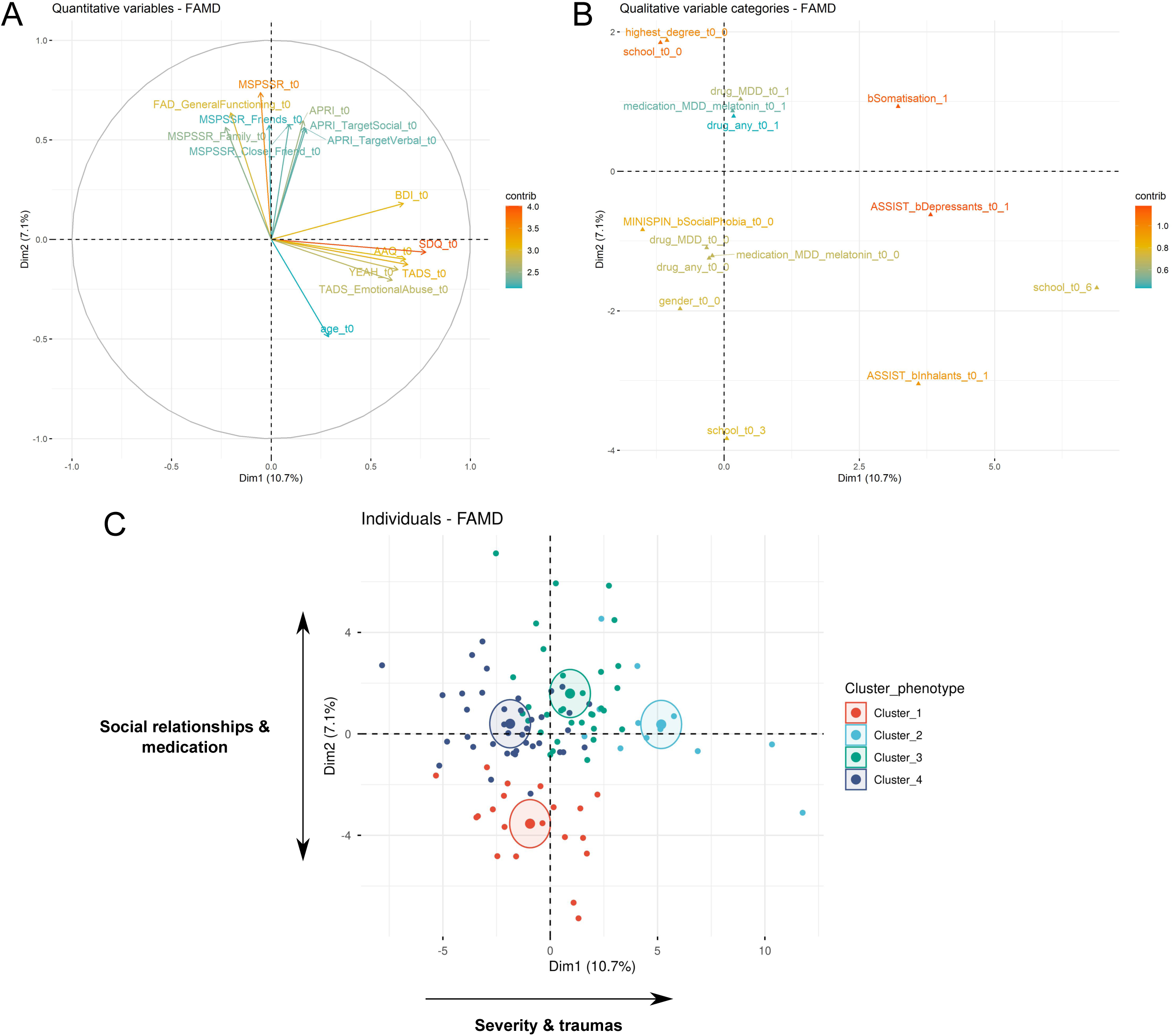
Clinical phenotypes of adolescent depression. Top 15 A) quantitative and B) qualitative drivers for the clinical phenotypes in Factor Analysis for Mixed Data, and C) phenotype clusters of 103 adolescents with depression using K-means clustering. Colored ellipses represent cluster centers. MSPSSR = Multidimensional Scale of Perceived Social Support, FAD = Family Assessment Device, APRI = Adolescent Peer Relations Instrument, BDI = Beck Depression Inventory, SDQ = Strengths and Difficulties Questionnaire, AAQ = Acceptance and Action Questionnaire, YEAH = Youth Experiences and Health, TADS = Trauma and Distress Scale, ASSIST = the Alcohol, Smoking and Substance Involvement Screening Test, MINISPIN = Mini Social Phobia Inventory, MDD = Major Depressive Disorder, t0 = baseline questionnaire. Categorical binary variables coded as 0 (no) and 1 (yes), school 0/3/6 = comprehensive school/university of applied sciences/unclear or undefined, highest_degree_t0_0 = part of comprehensive school.

Four phenotype clusters were generated, mostly reflecting the first two dimensions driven by severity and traumas (Dim1), and social relationships and medication (Dim2) (Fig. 3C). Cluster 1 (n=20) included a relatively higher proportion (35%) of boys, the lowest level of perceived social support, comorbidities, and medication. Cluster 2 (n=12) was characterized by the most severe, trauma-related depression with multi-comorbidity (mostly with somatization, panic, and psychotic disorders), the most severe insomnia, and a high level of smoking (90%) and substance use. All individuals in cluster 2 had MDD, self-mutilation, and alcohol drinking. Cluster 3 (n=32) included individuals who were most often medicated (75%), had dysthymia and double depression (> 60%), and commonly reported self-mutilation (87 %). Experiences of bullying victimization were common in clusters 2 and 3. Cluster 4 (n=39) indicated a less severe depressive phenotype with the lowest scores for social phobia, peer problems, and internet addiction; however, it showed a relatively higher level of divorced parents (61%), smoking (70%), cannabis (21%), and alcohol (87%) use. Details of the cluster-specific features are presented in Table A.8.

### 3.5. Correlations between proteomic and phenotype profiles

Nominally significant correlations were found between baseline proteomic and phenotypic profiles of 46 adolescents with DD in specific components (Fig. B.4, Table A.9); namely PC3 and Dim2 (r_s_ = -0.33, p = 0.026), and PC4-5 and Dim3 (r_s_ = -0.30, p = 0.043; r_s_ = -0.32, p = 0.032). Phenotypic dimension 2, capturing social relationships and medication, was linked to proteins involved in the regulation of complement and coagulation cascades, and lipoprotein particles (Table A.10a). Dim3 was specifically driven by experiences of bullying victimization and peer problems (Fig. B.5) and associated with proteins involved in ECM-receptor interactions, ECM proteoglycans, complement and coagulation cascades, and PI3K-Akt signaling pathway (Table A.10b).

### 3.6. Changes in proteins and symptoms over 6-month follow-up

Regression models using 40 paired samples identified alterations in 109 nominally significant (p < 0.05) proteins overall, 146 proteins in 36 girls, and 19 in 4 boys (Table A.11a). However, none of these remained significant after multiple testing. The altered proteins were involved in biological processes such as axon development, cell morphogenesis, and cell adhesion. In girls, proteins involved in platelet aggregation and glycosaminoglycan metabolic processes were also identified. Additionally, enrichment was observed for cellular components such as cell-substrate junctions and collagen-containing extracellular matrix (Table A.11b).

Overall, significant (adj. p < 0.05) improvements were observed among 75 adolescents with DD in mental well-being, especially in depressive, emotional and insomnia symptoms during the 6-month follow-up (see Fig. 4, Fig. B.6, and Table A.12). The largest improvements were seen in phenotype clusters 2 and 3, showing the most severe depression at baseline. The symptom scales showing significant changes over the follow-up were used to identify longitudinal protein-symptom associations in 40 adolescents. The strongest associations between protein and symptom changes were seen for BDI, showing significant (p < 0.05) associations with 128 protein changes (Table A.13a).

**Figure 4.**
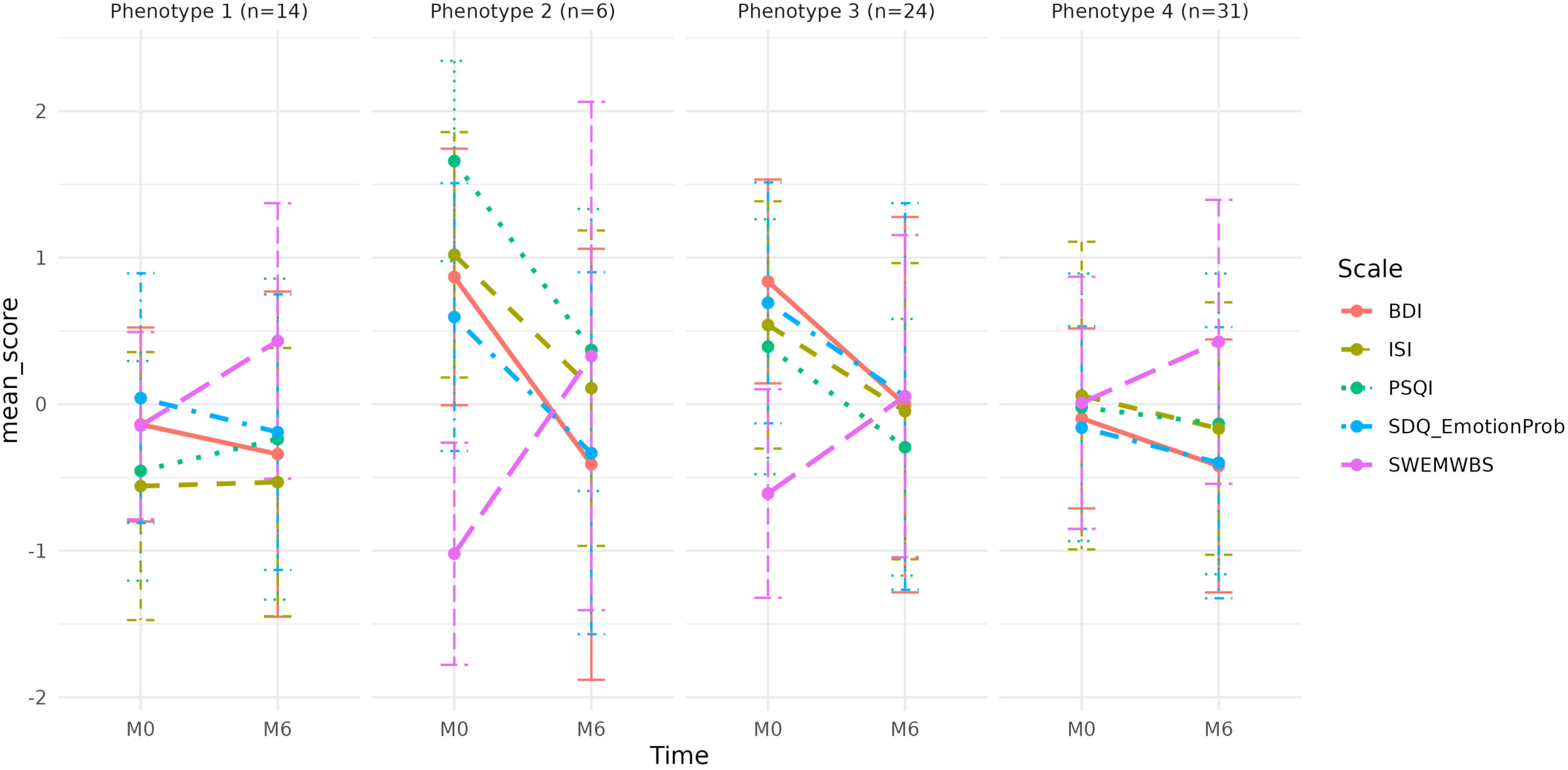
Standardized changes in symptom scores over the 6-month follow-up in adolescents with depression. Visualization of the balanced data for 75 adolescents. Significant scales among 59 adolescents with DD in a linear mixed-effect model adjusted for age, sex and DD-medication over the 6-month follow-up, Benjamini-Hochberg-adjusted p-value < 0.05. BDI = Beck Depression Inventory, PSQI = Pittsburgh Sleep Quality Index, ISI = Insomnia Severity Index, SDQ_EmotionProb = Strengths and Difficulties Questionnaire/emotional problems-subitem, and SWEMWBS = Short Warwick-Edinburgh Mental Wellbeing Scale. © NHS Health Scotland, University of Warwick and University of Edinburgh, 2008, all rights reserved.

These proteins were involved, e.g. in collagen-containing extracellular matrix and basement membrane, granules of cytoplasmic vesicles and endomembrane system, complement and coagulation cascades, and protein-lipid complexes (Table A.13b). However, none of these remained significant after FDR correction.

## 4. Discussion

We identified alterations in the plasma proteome between depressed and healthy adolescents, suggesting changes in stress-response-related pathways in adolescent depression. These alterations included dysfunction of the immune system and hemostasis, particularly decreased complement proteins, glucose metabolism, intracellular signaling pathways, and increased proteasome components related to oxidative stress. Furthermore, adolescents with depression exhibited reduced levels of proteins involved in extracellular matrix organization and keratan sulfate biosynthesis.

Psychological stress-induced inflammation is a key risk factor for depression in adults, especially in individuals with childhood adversity and trauma (X. Feng et al., 2025). The abundance of C-reactive protein (CRP), the most common marker of low-grade inflammation, was higher in girls with depression in our study, but no differences were seen in interleukin-6 (IL-6). Additionally, we observed an increased abundance of tumor necrosis factor (TNF) receptor components, binding pro-inflammatory interleukins, and TNF-α. Although adolescents with depression reported a wide range of traumatic experiences, peripheral pro-inflammatory responses were mainly limited to CRP and TNF-receptors. Thus, our findings suggest that systemic inflammation may play a smaller role in adolescent depression compared to adults.

We observed major changes in the complement cascade in adolescents with depression, who exhibited an overall decrease in complement proteins. Particularly, C8 and C9 components of the membrane attack complex and C3 were decreased in adolescents with depression. Furthermore, we observed a potential link between adolescent depression and autoimmune responses via complement components. Previous preclinical and clinical studies in adults have indicated the role of the complement cascade in depression; however, suggesting its activation and increased levels of C3 (T. Feng et al., 2020; Luo et al., 2022). On the other hand, immune system dysregulation, including immune activation and immune suppression, has been implicated in depression, yet the interconnection remains unclear (Blume et al., 2011; Maes & Carvalho, 2018). Besides the activated immune-inflammatory response system (IRS) in depression, the compensatory immune-regulatory reflex system (CIRS) has been introduced in past studies (Maes & Carvalho, 2018). The co-activation of CIRS dampens immune-inflammatory responses and can attenuate systemic pro-inflammatory effects (Maes & Carvalho, 2018). Furthermore, immune suppression impacts innate and adaptive immunity, for instance, by reducing the proliferation and activity of natural killer (NK) and T cells (Zorrilla et al., 2001). Additionally, parallel immune suppression and immune activation induced by persistent systemic inflammation might be an underlying mechanism connecting depression with autoimmune conditions, showing a high co-occurrence with psychiatric diseases (Mudra Rakshasa-Loots et al., 2025). Together, our findings with the current evidence suggest the role of immunosuppressive regulation and the IRS/CIRS balance, particularly in adolescent depression.

We identified increased abundances of proteins involved in the KEAP1-NFE2L2 (i.e. NRF2) pathway in adolescents with depression, suggesting that the body strives to adapt to oxidative stress. Further supporting this observation, we found increased NRF2-regulated proteins involved in reactive oxygen species (ROS) detoxification, such as superoxide dismutases, glutathione reductase, and glutathione peroxidase, in depression. Additionally, adolescents with depression reported several psychosocial and physiological stressors, such as trauma experiences, problems in relationships with family and friends, and sleep problems, particularly insomnia. Along with immune system dysregulation, oxidative stress is implicated in the pathophysiology of depression (Kuwar et al., 2025; Maes et al., 2025; Palta et al., 2014) and linked to both childhood adversity and internalizing symptoms in adolescents (Horn et al., 2019). Psychosocial and physiological stress activate the hypothalamic-pituitary-adrenal axis, which can induce oxidative stress (Aschbacher et al., 2013). This further contributes to cellular responses such as bioenergetic shifts and mitochondrial dysfunction, misfolded and damaged proteins, and activation of intracellular signaling cascades (Abdelwahed et al., 2022; Kuwar et al., 2025; Maes et al., 2025; Picard et al., 2015). Furthermore, oxidative stress and biomolecular damage are suggested as mediators for the effects of sleep deficiency, leading to persistent activation of stress responses (Coulson et al., 2022). ROS, produced by oxidative stress, activate antioxidant defense responses, primarily the KEAP1-NFE2L2 pathway, which attempts to restore redox balance. However, NRF2 signaling appears insufficient in depression, permitting persistent stress kinase activity and neuroimmune activation (Abdelwahed et al., 2022). While impaired NRF2 signaling and a decreased antioxidant buffer have been reported in depressed adults (Palta et al., 2014; Zuo et al., 2022), adolescents might retain stronger responses and antioxidant capacity. This can also contribute to the lower pro-inflammatory responses compared to adult depression.

Increased proteasome subunits observed in depressed adolescents were involved in several enriched pathways in the present study, including the KEAP1-NFE2L2 pathway, MAPK signaling pathway, and cell cycle regulation (e.g. G2/M checkpoint). Oxidative stress increases the load of damaged proteins and activates the ubiquitin–proteasome system (UPS), which shows an emerging role in mood disorders (Belaish et al., 2021; Santos et al., 2023). UPS serves as the key cellular machinery for protein degradation in proteasomes (Santos et al., 2023). In line with our findings, previous studies have reported upregulated proteasome subunit genes (Belaish et al., 2021) and proteasome system dysregulation (Frye et al., 2015) in MDD.

We identified alterations in MAPK and IGF-related signaling pathways in depression. The MAPK signalling pathway is an essential kinase family that regulates cell survival, growth, and neuroplasticity (Volmat & Pouysségur, 2001). In parallel with the antioxidant defense system, it is activated by ROS and regulates Nrf2 expression (P. Wang et al., 2023). Previous studies have provided mechanistic evidence linking the extracellular signal-regulated kinase (ERK) subfamily of the MAPK pathway to both the pathophysiology of depression and therapeutic antidepressant responses (Duman et al., 2007; J. Q. Wang & Mao, 2019). Downregulated ERK signaling in the central brain areas in depression has been reported in humans and animals (J. Q. Wang & Mao, 2019). We identified the opposite change in peripheral ERK signaling, mostly linked to its upstream activators (BRAF, RAF1) and increased proteasome components. Additionally, we found a decrease in proteins involved in the regulation and transport of IGFs, such as insulin-like growth factor binding proteins (IGFBP) 3 and 5. Although the previous studies have indicated altered peripheral IGF signaling in depression, the decrease in IGFBP-3 and IGFBP-5 might also be a response to antidepressant treatment (Fernández-Pereira et al., 2023; Whipp et al., 2025). Overall, these partly contradictory findings might reflect a peripheral compensatory mechanism or enhanced regulatory responses of cell proliferation and survival under increased cellular stress in depressed adolescents.

In this study, adolescents with depression exhibited increased enzymatic proteins involved in energy-producing glucose metabolism. Mitochondrial energy production is another essential cellular mechanism for stress adaptation (Picard et al., 2015). Past studies have demonstrated shifts in energy metabolism in depression in both the brain (Su et al., 2014) and blood (Chen et al., 2024). Interestingly, activated gluconeogenesis and metabolic enzymes in all energy-producing pathways were observed in a depressive-like animal model, potentially compensating for reduced citric acid cycle activity due to mitochondrial dysfunction (Ling-Hu et al., 2021). Our findings align with the hypothesis of protein-level adaptation to energy deficit caused by stress-related mitochondrial dysfunction. ROS-driven oxidative stress impairs mitochondrial function, while MAPK and NRF2 signaling act to preserve redox homeostasis, indirectly connecting redox regulation to energy metabolism pathways. However, under conditions of prolonged stress, these cellular adaptive mechanisms may become insufficient, thereby contributing to pathophysiological progression. Together, our findings highlight the role of oxidative stress-induced systemic adaptation in adolescent depression.

Furthermore, we observed alterations in extracellular matrix (ECM) proteins and keratan sulfate biosynthesis in the plasma of depressed adolescents, suggesting altered tissue remodeling processes. Keratan sulfate proteoglycans are structural components of ECM in several tissues, but also in the nervous system (Caterson & Melrose, 2018). Interestingly, ECM-related proteins were linked to symptom dimensions of social well-being and a change in depressive symptoms during the follow-up. However, these associations did not remain significant after FDR correction. Previous experimental studies have demonstrated that particularly social stress induces changes in the brain’s ECM network, such as altered expression of ECM and cell-cell interaction proteins and disturbances in glutamatergic synapses in the prefrontal cortex and altered neuronal plasticity (Blanco & Conant, 2020; Koskinen et al., 2020; Zhang et al., 2024). Although our results suggest involvement of peripheral ECM composition in adolescent depression, it is not yet clear whether these observations relate to the brain ECM changes or to identified psychosocial factors.

Regarding the role of social well-being in adolescent depression, we identified traumatic experiences and social relationships together with symptom severity as the strongest nominators for the phenotypic clusters. However, the clusters were not clearly distinct from each other; particularly, clusters 3 and 4 showed notable overlap. Bullying victimization and sexual, physical, and emotional abuse were observed as common factors among adolescents who showed the most severe phenotype. On the other hand, low perceived social support and the second-highest average scores for emotional neglect and abuse were observed in cluster 1, suggesting a lack of social and emotional connection. Nearly all adolescents with depression reported problems in their family relationships. Childhood adversities and trauma are well-characterized risk factors for depression (Alisic et al., 2019; Heim & Nemeroff, 2001) and have been associated with poorer recovery from depressive symptoms (Sarnola et al., 2025). Altogether, these findings support the importance of a physically and emotionally safe and supportive living environment in adolescents’ mental health.

### Strengths and Limitations

A key strength of this study is the use of adolescent samples and unbiased shotgun proteomics, providing a snapshot of the body’s dynamic state. Proteomics profiles were determined at two points to identify protein associations with fluctuations in clinical symptoms. Additionally, phenotypic subtypes were defined to examine the heterogeneous manifestations of depression and their potential reflections in plasma proteomics. Healthy controls in the PANIC study had high psychological well-being, enabling a strict selection of control samples without depressive-like symptoms or other confounding factors.

The number of statistically significant differences in proteins and the effect sizes between DD and HC were notably high. We acknowledge that the extent of the findings may be biased due to the inclusion of two independent cohort studies, which introduce a non-adjustable cohort-driven batch effect. Although the cohorts were similar in age, geographical location, and blood sampling protocols, differences in sample processing temperature and freezing may have affected the abundance and detection of certain proteins, particularly those involved in coagulation pathways (Geyer et al., 2019). Most of the coagulation panel quality markers suggested by Geyer et al. were among the proteins that differed significantly between the groups (Geyer et al., 2019). However, the platelet-related and hemostatic pathways were not composed solely of these potential contaminants. Additionally, residual effects of medications in the DD group may persist and confound the results. Due to the expected cohort-driven batch effect, a cohort-specific analysis was performed for DD samples, and a stricter protein selection was used for enrichment analysis.

While girls showed generally stronger and wider-ranging biological alterations, interpretations of the sex-specific differences should be avoided, given the unbalanced sex distribution. There was a higher proportion of girls in the DD group than in HCs. This difference may introduce bias, as certain proteomic differences are sex-specific, and boys were statistically underpowered. To minimize this bias, the linear regression models were adjusted for sex, and the sex-specific sensitivity subgroup analyses were performed. Further investigation is needed to reliably characterize sex-specific differences. Although the plasma proteome is influenced by body composition, body mass index (BMI) was excluded from the main analysis due to insufficient data on body weight or height and expected non-random missingness. A sensitivity analysis including a subset of adolescents with BMI data suggested that most observed protein associations were not primarily influenced by BMI.

Finally, longitudinal analyses revealed only nominally significant changes in proteins, which may be due to the relatively small number of individuals and the short follow-up. As observed in this study, similar proteomic profiles were detected in adolescents independent of baseline symptom severity. This suggests that plasma proteomics alone is insufficient to account for the heterogeneity in subjective symptom experiences, and more comprehensive body-brain approaches are needed.

## 5. Conclusions

This study demonstrates plasma proteomic alterations in adolescents with depression, characterized by immune dysregulation, redox–proteostasis adaptation to oxidative stress, and extracellular matrix remodeling. These biological processes illustrate the systemic adaptation to psychosocial stress. Our findings suggest the need for integrative, context-informed approaches that bridge immune-metabolic, redox-proteostasis, and psychosocial domains to advance mechanistically driven interventions in youth depression.

## Supporting information

Appenedix B_Supplementary Figures

## CRediT authorship contribution statement

**Aino-Kaisa Piironen**: Conceptualization, Formal analysis, Visualization, Writing - original draft, Writing - review & editing, Funding acquisition, Project administration. **Alexey M. Afonin**: Methodology, Writing - review & editing. **Karoliina Kurkinen**: Funding acquisition, Writing - review & editing. **Timo A. Lakka**: Resources, Conceptualization, Supervision, Writing - review & editing. **Tommi Tolmunen**: Resources, Conceptualization, Funding acquisition, Supervision, Writing - review & editing. **Katja M. Kanninen**: Supervision, Funding acquisition, Conceptualization, Resources, Project administration, Writing review & editing.

## Funding

This project has received funding from the European Union’s Horizon 2020 research and innovation programme under grant agreement No 874724. Equal-Life is part of the European Human Exposome Network. Researcher Piironen has received personal grants from the Finnish Cultural Foundation, Yrjö Jahnsson Foundation, and the Finnish Brain Foundation. Researcher Kurkinen has received grants from the Finnish Brain Foundation and the University of Eastern Finland, Institute of Clinical Medicine. Researchers Tolmunen and Piironen were supported by the Strategic Research Council within the Research Council of Finland (SchoolWell, grant number 372260, work package 372266).

## Acknowledgments

We thank all youth and their caregivers who participated in the SMART and PANIC studies, the research nurses Kaisa Korhonen and Kirsi Saastamoinen, and the Eastern Finland Laboratory Centre Joint Authority Enterprise, for sample and data collection. We acknowledge the Turku Proteomics Facility, supported by Biocenter Finland, for performing mass spectrometry analyses, Tero Sievänen from the UEF Bioinformatics Center, Biocenter Kuopio, Biocenter Finland, University of Eastern Finland, Finland, for consulting, and the laboratory assistance of Mirka Tikkanen. The computational analyses were performed on servers provided by UEF Bioinformatics Center, Biocenter Kuopio, Biocenter Finland, University of Eastern Finland, Finland.

## Declaration of competing interests

The authors declare no competing interests.

## Data availability

The data contain personal information of adolescents participating in SMART and PANIC studies. According to ethical approvals, these data should be kept confidential. Data are available from the corresponding author and/or cohort owners upon reasonable requests for researchers who meet the criteria for access to confidential data. A data-use and transfer agreement is required to protect privacy and ensure compliance with national data protection legislation, the content and specific clauses of which will depend on the nature of the requested data.

## Notes

### Competing Interest Statement

The authors have declared no competing interest.

### Clinical Trial

NCT01803776, NCT06262958

### Author Declarations

The Research Ethics Committee of the Hospital District of Northern Savo approved the SMART study (238/2017) and the PANIC study before baseline in childhood (69/2006) and before the 8-year follow-up in adolescence (422/2015). All adolescents and caregivers of those younger than 15 years old in the SMART study provided written informed consent. Adolescents in the 8-year follow-up of the PANIC study also provided written informed consent.

### Summary of Updates

Additional sensitivity analyses were performed to consider potential confounding effects of alcohol use, smoking, and body mass index; Sections 2.7.5. Sensitivity analysis, 3.2. Plasma protein alterations in depression, Strengths and Limitations, and Supplementary files updated.

