## Supplementary material for "Untargeted plasma proteomics and clinical phenotypes in adolescent depression": Appenedix B_Supplementary Figures

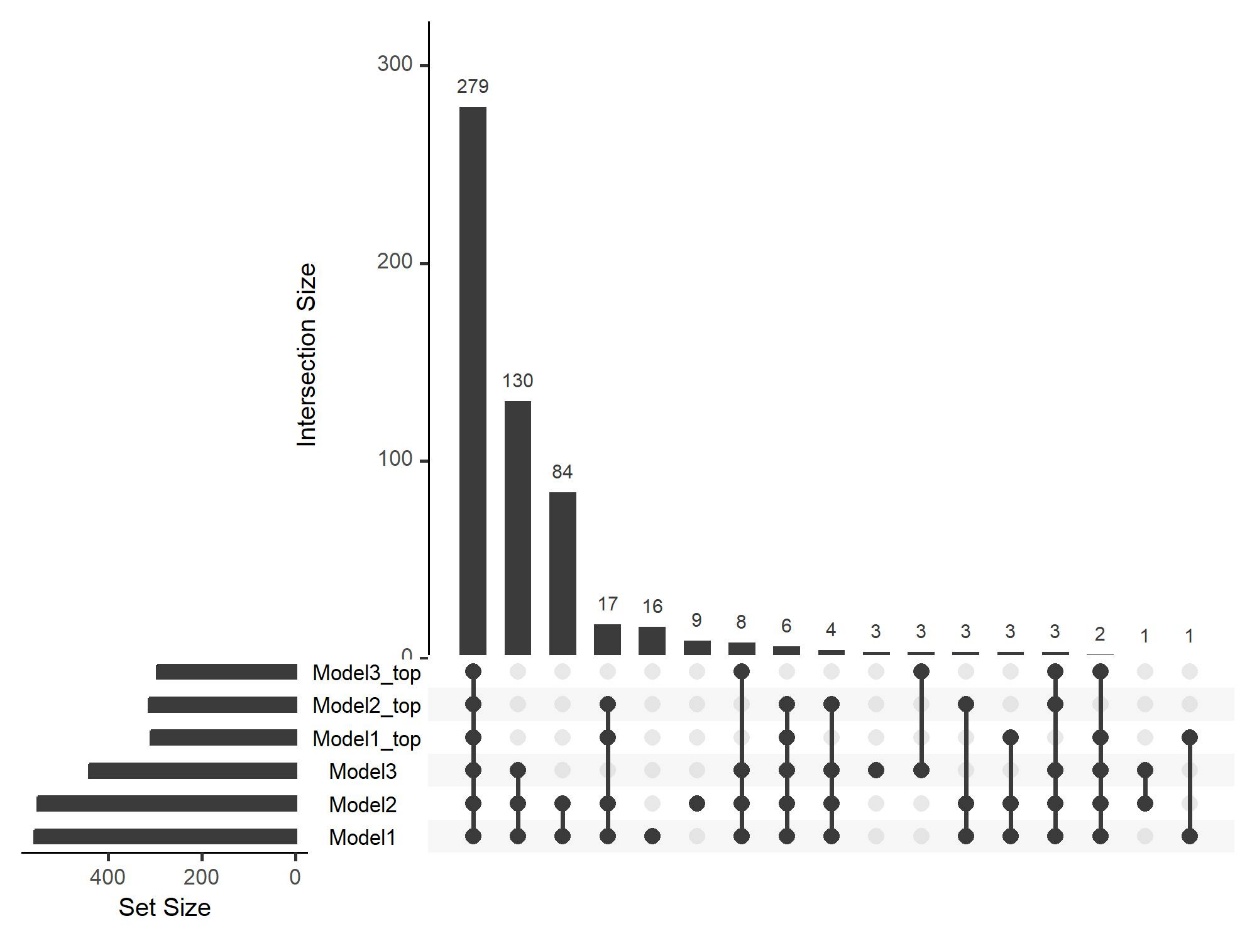

**Figure B.1. UpSet Plot on significant proteins in linear regression models.** Comparisons of significantly different proteins between adolescents with depression at baseline and healthy controls. Model 1 was adjusted for age, sex, and medication use; model 2 was adjusted for age, sex, medication use, smoking, and alcohol use; and model 3 was adjusted for age, sex, medication use, and body mass index. Protein sets of models 1-3 include proteins with adj. p < 0.05, and models 1-3_top include proteins with adj. p < 0.01 and the absolute Log2FC ≥ 1.

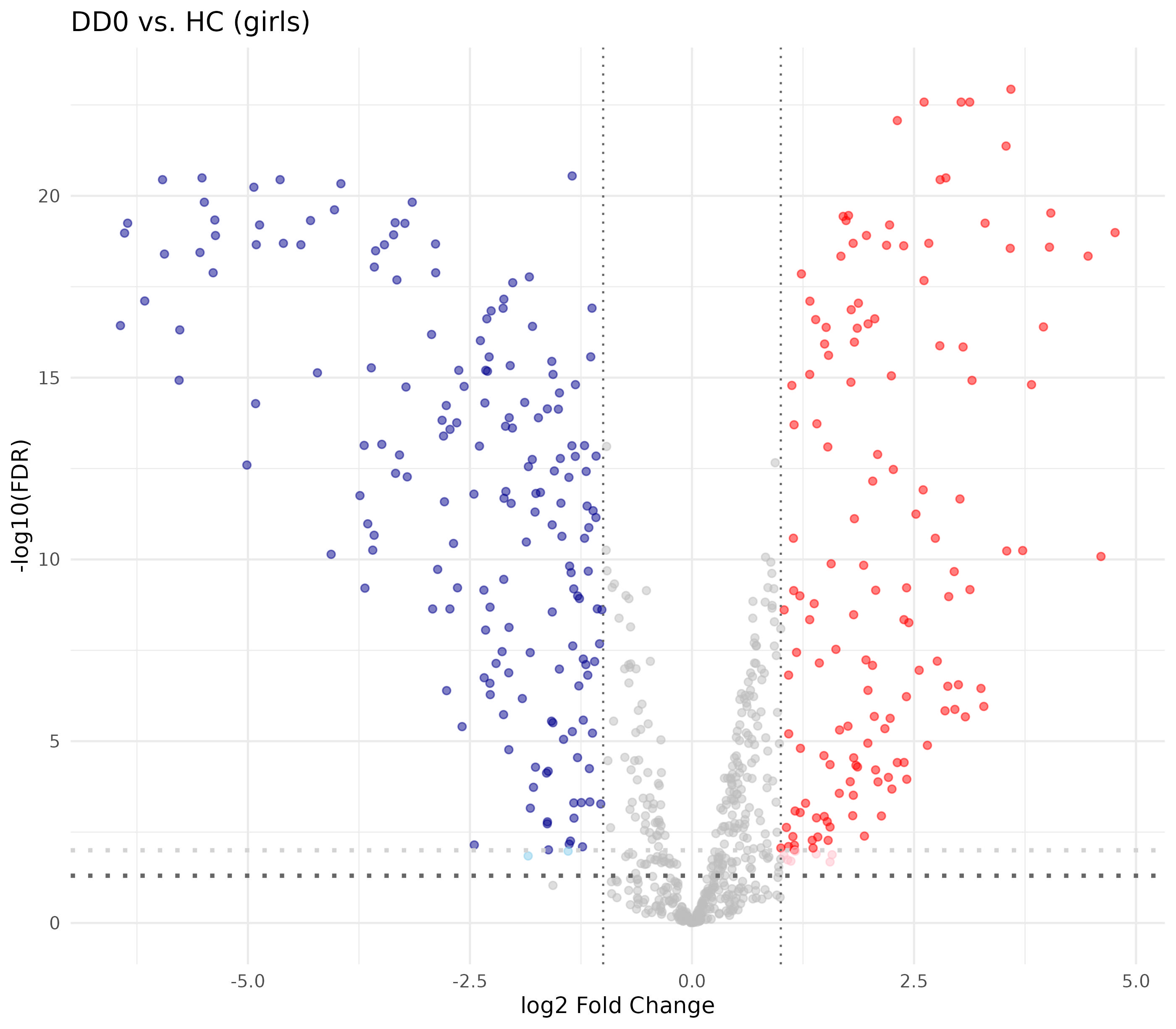

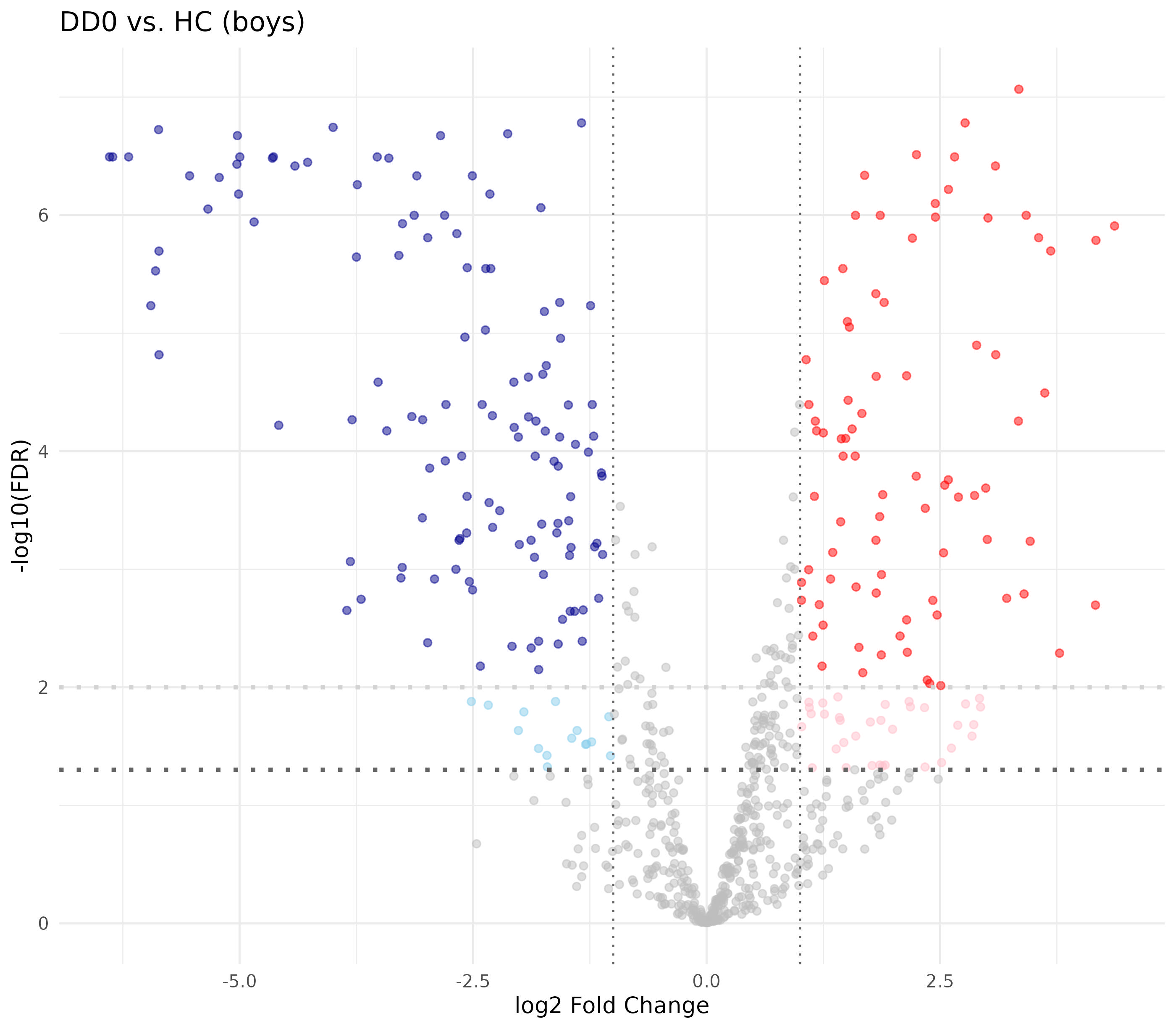

**Figure B.2. Differentially abundant proteins in sex-specific analysis for depression vs HC.** Proteins decreased in adolescents with depressive disorders (DD) compared to healthy controls (HC) are shown in blue, and proteins increased in red. In girls (top, nDD=42, nHC=24), 313 proteins and 207 proteins in boys (bottom, nDD=5, nHC=25) with adj. p < 0.01 and the absolute Log2FC ≥ 1. Light blue and red indicate additional proteins with adj. p = 0.01-0.05.

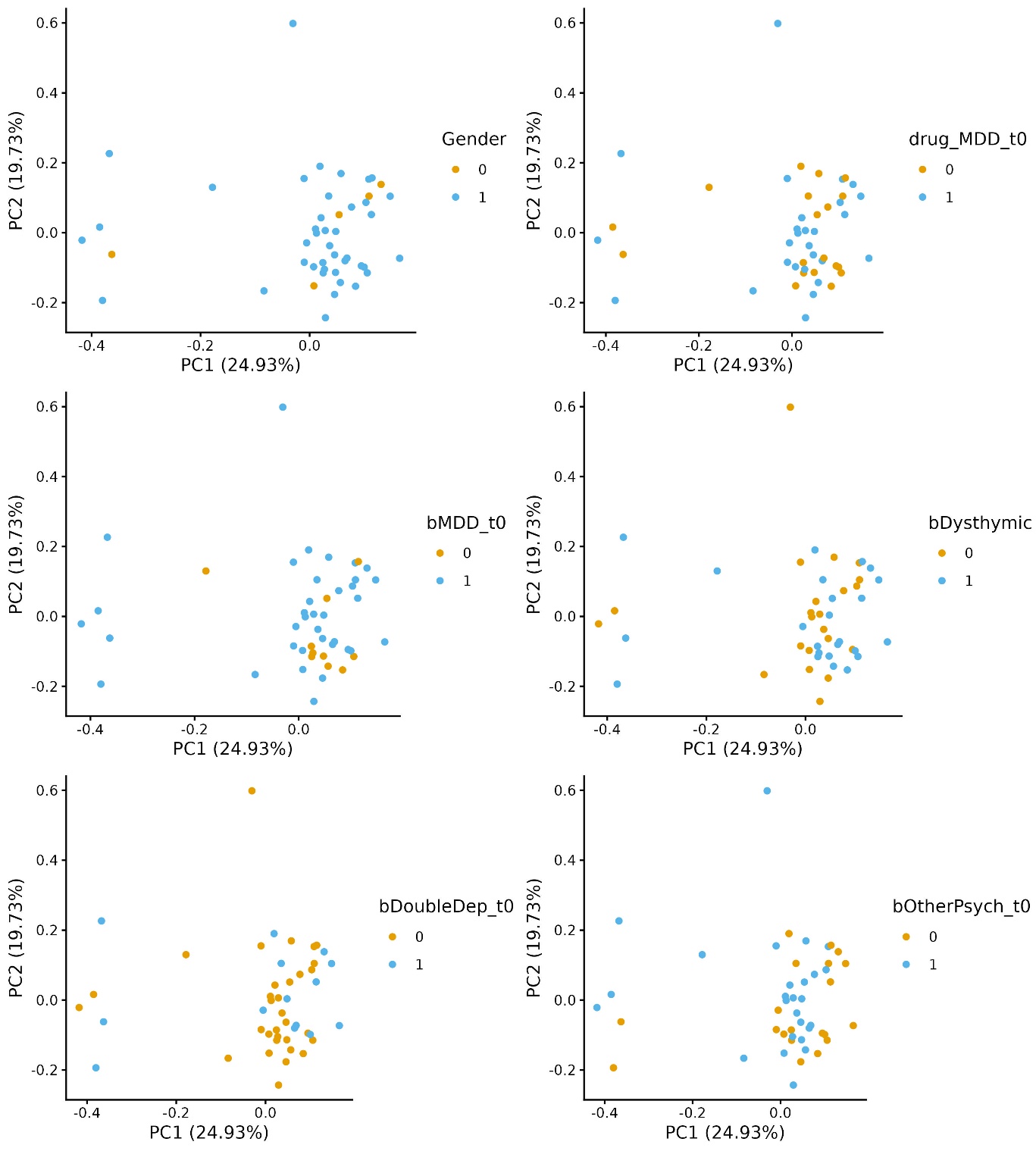

Sex

**Figure B.3.** **Proteomic profiles in depression by sex, medication and sub-diagnoses.** Adolescents with depression at baseline (n=46). MDD = major depressive disorder, Dysthymic = dysthymic disorder, DoubleDep = MDD and dysthymic disorder, OtherPsych = other psychiatric disorders, Gender 0 = boy, 1 = girl. Otherwise, 1 = yes, 0 = no.

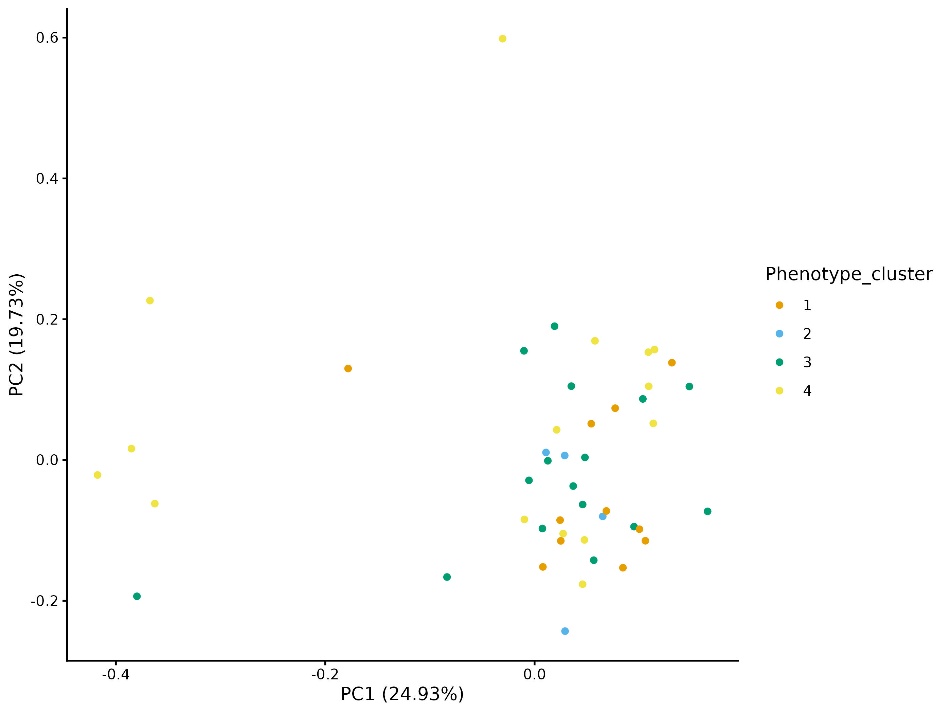

**B**

**A**

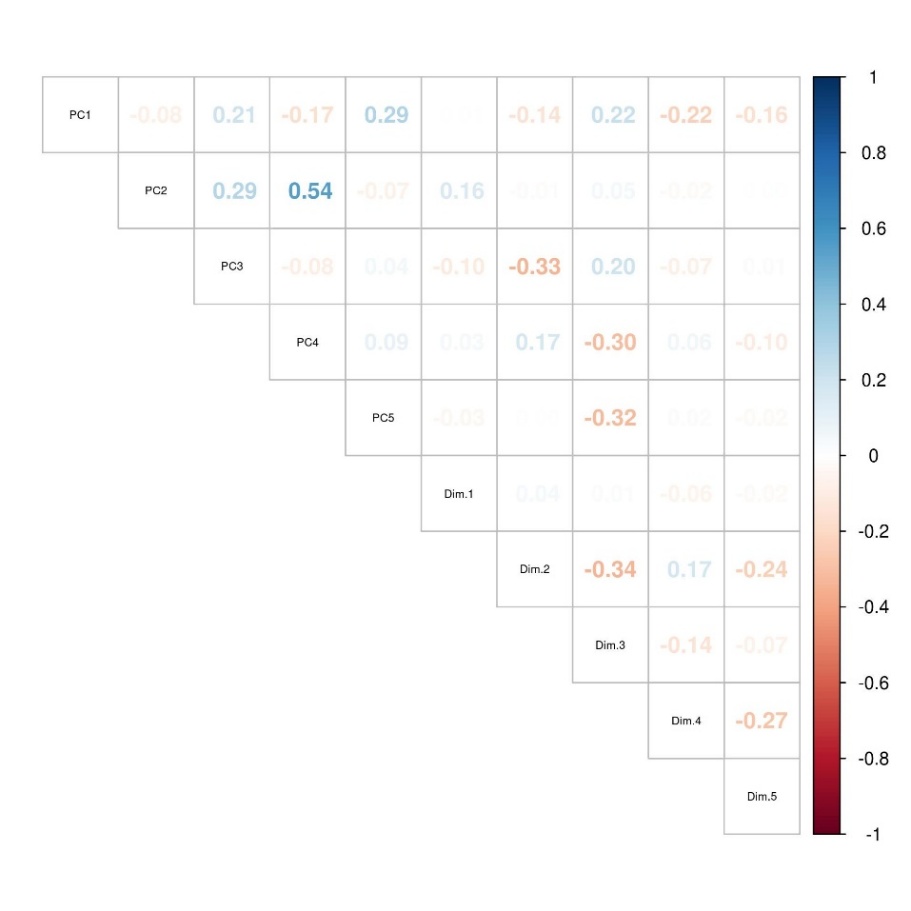

**Figure B.4. Correlations between proteomics and phenotype profiles. A)** PCA on proteomics profiles in adolescents with depression at baseline (n=46), colored by phenotype clusters. **B)** Correlation coefficients for PC and FAMD dimensions. Two-sided Spearman’s pairwise correlation.

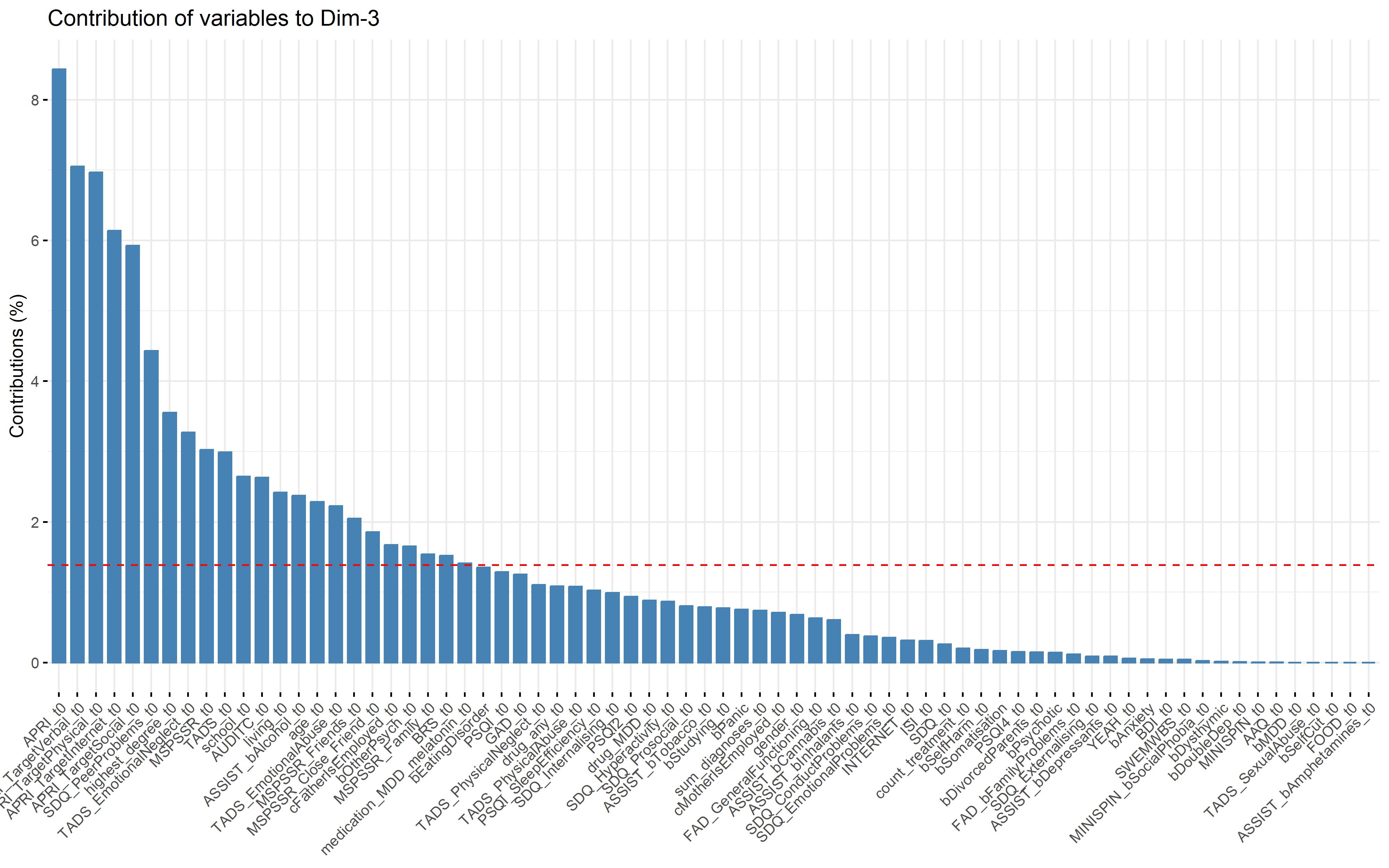
**Figure B.5. Contributions of clinical variables to the third dimension in the Factor Analysis for Mixed Data.** Adolescents with depression at baseline (n=103).

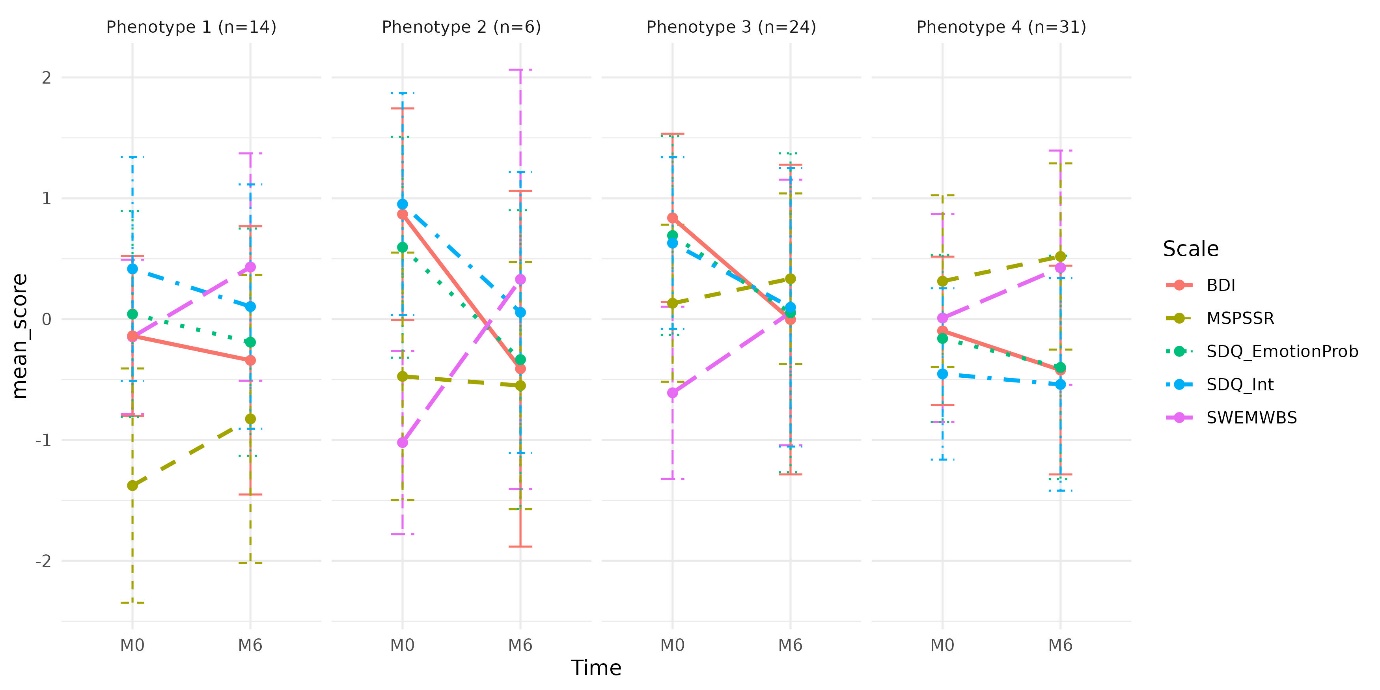

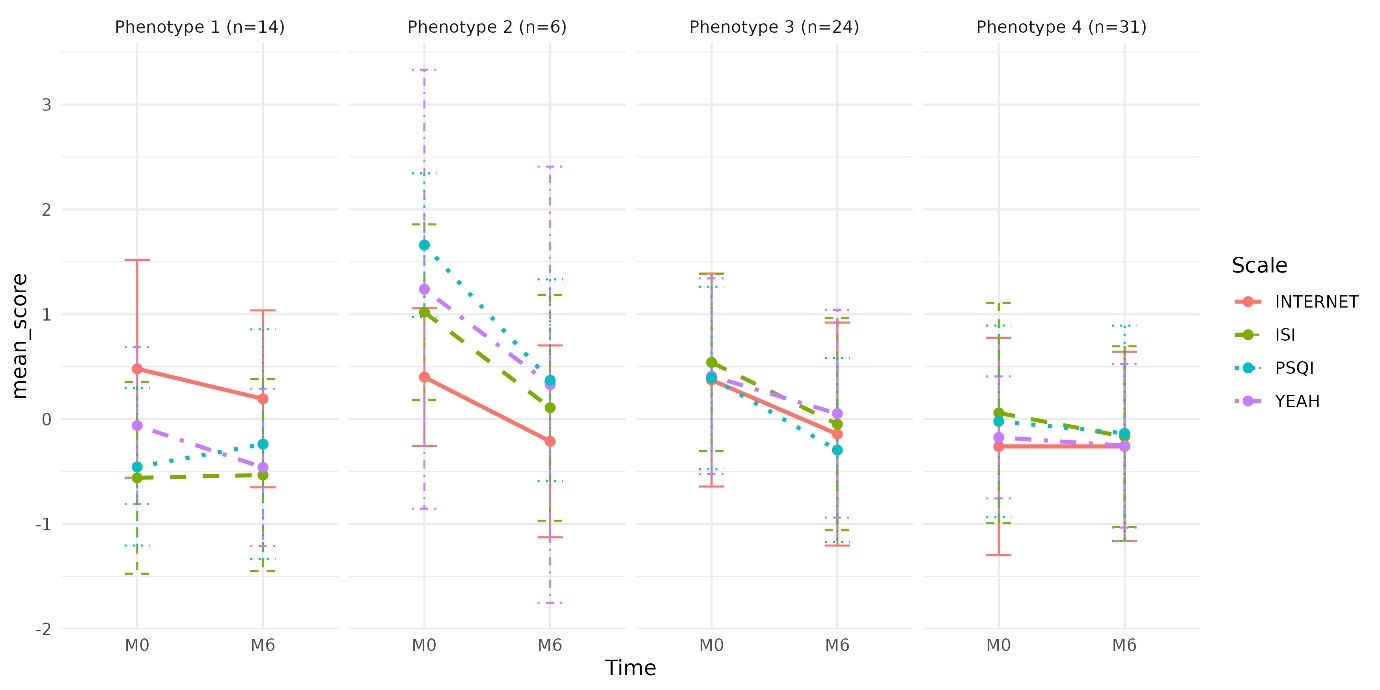

**Figure B.6. Standardized changes in symptom scores over the 6-month follow-up in adolescents with depression.** Significant scales (adj. p < 0.05) in a linear mixed-effect model adjusted for age, sex and baseline DD-medication using balanced data for 75 adolescents. **A)** BDI = Beck Depression Inventory, MSPSSR = Multidimensional Scale of Perceived Social Support, SDQ_EmotionProb = Strengths and Difficulties Questionnaire/emotional problems, SDQ_int = Strengths and Difficulties Questionnaire/internalizing symptoms, SWEMWBS = Short Warwick-Edinburgh Mental Wellbeing Scale (© NHS Health Scotland, University of Warwick and University of Edinburgh, 2008, all rights reserved) and **B)** INTERNET = Internet Addiction Test, ISI = Insomnia Severity Index, PSQI = Pittsburgh Sleep Quality Index, YEAH = Youth Experiences and Health. M0 = baseline, M6 = 6-month follow-up.
